# A sagittal gradient of pathological and compensatory effects of neurophysiological slowing in Parkinson’s disease

**DOI:** 10.1101/2022.08.05.22278436

**Authors:** Alex I. Wiesman, Jason da Silva Castanheira, Clotilde Degroot, Edward A. Fon, Sylvain Baillet, PREVENT-AD Research Group, Quebec Parkinson Network

## Abstract

Using magnetoencephalographic imaging and extensive clinical and neuropsychological assessments, we show that patients with Parkinson’s disease (PD; N = 79) exhibit a slowing of neurophysiological activity relative to healthy adults (N = 65), which relates to motor and cognitive abilities. Importantly, the association between neurophysiological slowing and PD clinical features varies systematically across the cortex along a sagittal gradient: cortical slowing is associated with worse impairment in dorsal-posterior cortices, and this association is reversed in ventral-anterior cortical regions. This pathological-to- compensatory anatomical gradient is sensitive to differences in patients’ individual clinical profiles, and co- localizes with normative atlases of neurotransmitter receptor/transporter density. Long-range functional connectivity between posterior regions and parietal and frontal cortices is also significantly shifted towards lower frequencies in PD, demonstrating a novel network-level slowing effect. Taken together, these findings demonstrate the multifaceted nature of neurophysiological slowing in patients with PD, with anatomically-dependent clinical relevance to motor and cognitive symptoms.

## Introduction

Parkinson’s disease (PD) is the second most common neurodegenerative disorder worldwide^1^. It is characterized by hallmark declines in motor functions^2^, with many patients also experiencing debilitating declines in cognitive abilities^3^. Although the etiology of PD is not clear to date, the neuropathological process includes a progressive degeneration of dopaminergic neurons and glial cells in the substantia nigra pars compacta, leading to dysfunctional dopamine (DA) signaling along the nigrostriatal pathway^2^. This leads to over-inhibition of thalamic projections to the cortex that are essential for the accurate execution of voluntary movements in the healthy brain^2^. Changes in cortical signaling are also well-documented in PD, yet the functional consequences of these cortical aberrations are not entirely clear. Some research has suggested that they convey compensatory effects (i.e., greater changes relative to healthy controls relating to better clinical outcomes) while others have instead indicated that they are deleterious^4-19^. These effects may not be mutually exclusive. Complex patterns of neurophysiological changes observed in patients with PD may simultaneously indicate both dysfunction and adaptive compensation, alongside additional effects of pharmacotherapies and clinical interventions intended to remediate PD symptoms. A more nuanced understanding of pathological versus compensatory effects of PD-related neurophysiological changes is needed to inform and advance interventions, such as targeted neuromodulation strategies to ameliorate symptoms and enhance compensatory capabilities in patients with PD^20-28^.

Neurophysiological indicators of PD pathophysiology include frequency-specific components of the rich spectrum of brain electrophysiology. Notably, beta-band (15 – 30 Hz) activity is hypersynchronous across the cortico-basal ganglia circuit in patients with PD^17,29-33^, relates to severity of motor dysfunction, and can be normalized by common therapeutics^34-38^. In the cortex of patients with PD, decades of electrophysiological studies have demonstrated a stereotyped pattern of frequency-defined neural changes relative to healthy adults, including both increased activity in low-frequency bands (e.g., delta [2 – 4 Hz] and theta [5 – 7 Hz]) and concurrent decreased power in high-frequency bands (e.g., alpha [8 – 12 Hz] and beta)^12,39-42^. This has led to a hypothesized *slowing* of brain activity in patients with PD, but it remains unclear whether such a neurophysiological effect relates to clinical features of the disease, and whether any such relationships are of a deleterious or compensatory nature. These multi-spectral deviations from healthy levels also comprise rhythmic and/or arrhythmic components^43-46^, for which clinical interpretation and significance for frequency-specific neuromodulation therapies remain to be established in PD^19,42,47-51^.

We adapted a recent measure of neurophysiological slowing^52^ (Figure 1A) with magnetoencephalography (MEG) data from a large sample of patients with PD (N = 79) and a matched group of healthy older adults (N = 65). We related neurophysiological slowing effects measured in the PD group to detailed clinical and neuropsychological indicators of motor and cognitive deficits, with the hypothesis that stronger cortical slowing is associated with greater clinical impairments. What we actually observed was that this association varied in a structured manner across the cortex, indicating a progressive change from compensation to impairment along the sagittal cortical plane (Figure 1B). To determine the clinical and neurochemical nature of this sagittal gradient effect, we investigate its sensitivity to clinical profile features and neurotransmitter receptor densities that are salient in PD. Finally, we provide evidence that cortical slowing also affects frequency-specific, inter-regional functional connectivity, which indicates a network-level slowing of neurophysiology in patients with PD.

**Figure 1.**
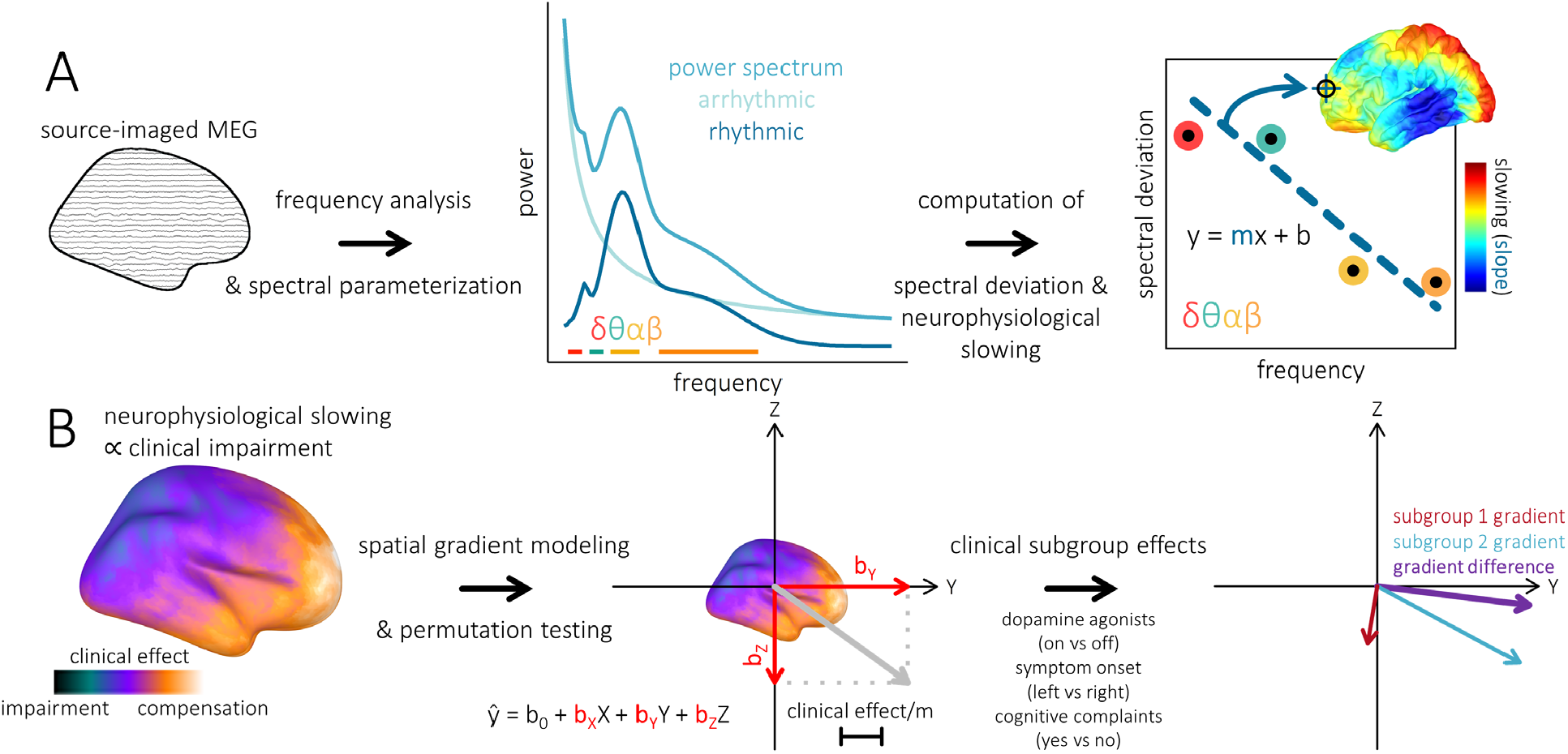
Neurophysiological slowing and anatomical gradient analyses. (A) Neural slowing computation. Source-imaged magnetoencephalography (MEG) data is first frequency-transformed and the vertex-wise power spectral densities (PSD) parameterized using *specparam*. The resulting PSDs are averaged over typical frequency bands (i.e., delta: 2–4 Hz; theta: 5–7 Hz; alpha: 8–12 Hz; beta: 15–29 Hz) and each spectrally- and spatially-resolved power estimate of neurophysiological signal per patient is normalized to the mean and standard deviation of the comparable estimates in the healthy control group. Within each patient and at each spatial location, a linear model is then fit to these spectral deviations across frequencies, and the slope of this model is extracted that represents the relative slowing (i.e., negative slope values) of brain activity relative to healthy levels. This procedure is performed per cortical vertex, resulting in a spatially-resolved map of neurophysiological slowing per patient. (B) Spatial gradient analysis. Cortical surfaces are first smoothed to reduce the impact of gyrification on the estimation of spatial gradient effects. Per each vertex location, neurophysiological slowing values are separately correlated with motor (i.e., UPDRS-III scores) and cognitive (i.e., sign-reversed neuropsychological scores averaged over cognitive domains) impairments, beyond the effects of age. These partial correlation maps are then linearly-scaled (i.e., using the Fisher-transform) and summed per vertex, resulting in a single cortical map showing the nature and strength of the relationships between neurophysiological slowing and clinical impairments across the brain. A linear multiple regression is then fit to these data and the beta weights extracted, with each of the cardinal axes (X: left – right; Y: posterior – anterior; Z: inferior – superior) represented as a predictor. The neurophysiological slowing data are then randomly permuted across patients and the partial correlation and spatial multiple regression steps repeated 1,000 times, with the resulting beta weights extracted and used to build null distributions per each predictor. To test for the effect of binary clinical factors on these gradients, the same procedure is performed within each binarized patient subgroup, with the difference in beta weights between the two subgroups used as the statistic of interest.

## Results

### Slowing of Rhythmic and Arrhythmic Neurophysiological Activity Relates Differentially to Clinical Impairments

Patients with Parkinson’s disease exhibited slowing of cortical neurophysiological activity, with the strongest effects in bilateral parieto-occipital cortices (TFCE; *p*_FWE_ < .001; peak vertex = x: -50, y: -77, z: 1; Figure 2A). The magnitude of this slowing effect was related to cognitive abilities (Figure 2B) including general cognitive function in left superior frontal cortex (TFCE; *p*_FWE_ = .048; peak vertex = x: -4, y: 63, z: 20), as well as domain-specific impairments in language in bilateral prefrontal and temporal regions (TFCE; *p*_FWE_ = .012; peak vertex = x: 4, y: 38, z: 11), attention in bilateral inferior frontal and somatomotor cortices (TFCE; *p*_FWE_ = .030; peak vertex = x: 50, y: -13, z: 52), and visuospatial function in bilateral anterior temporal regions (TFCE; *p*_FWE_ = .014; peak vertex = x: -49, y: -8, z: -44).

**Figure 2.**
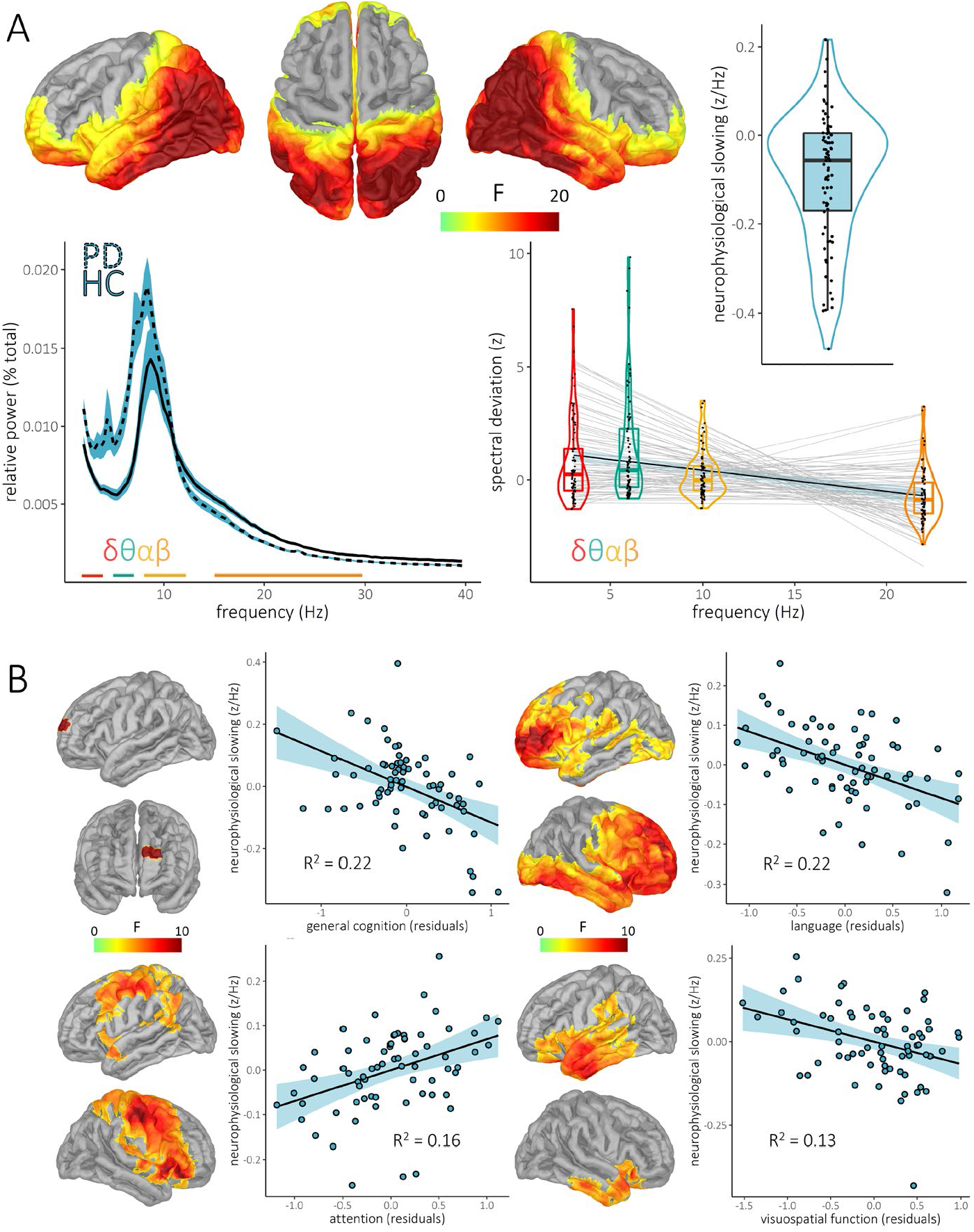
Neurophysiological slowing associated with cognitive abilities in Parkinson’s disease. (A) Cortical maps indicate significant clusters of neurophysiological slowing in patients with Parkinson’s disease (PD) after stringent multiple comparisons correction. Power spectra to the bottom left indicate the underlying data used to compute neurophysiological slowing from the cortical vertex exhibiting the strongest effect, with colored bars underneath showing the bandwidths of typical frequency-band definitions used for averaging. The plot to the bottom right shows the individual patient spectral deviations at this same cortical vertex for each frequency band, with the light grey lines-of-best fit indicating individual neurophysiological slowing slopes, and the overlaid black line and blue shaded area representing the overall group effect and 95% confidence intervals, respectively. These individual and mean neurophysiological slowing effects are also represented as single dots in the scatterplot to the top right. (B) Cortical maps indicate significant clusters where neurophysiological slowing was associated with cognitive function in patients with PD. Associated scatterplots indicate the nature and strength of this relationship at the cortical vertex exhibiting the strongest effect, with lines-of-best-fit, 95% confidence intervals, and *R*^*2*^ values overlaid.

Both arrhythmic (TFCE; *p*_FWE_ < .001; peak vertex = x: 45, y: -81, z: 7; Figure 3A) and rhythmic (TFCE; *p*_FWE_ < .001; peak vertex = x: 43, y: -71, z: 31; Figure 4A) neurophysiological generators contributed to the slowing effect. Arrhythmic slowing was stronger than rhythmic slowing in bilateral inferior frontal regions (TFCE; *p*_FWE_ = .018; peak vertex = x: 9, y: 40, z: -5; Figure S1), and no clusters were identified where rhythmic slowing was significantly stronger than arrhythmic. Arrhythmic cortical slowing was associated with motor impairments in bilateral prefrontal and temporal cortices (i.e., UPDRS-III scores; TFCE; *p*_FWE_ = .028; peak vertex = x: -41, y: 43, z: 19; Figure 3B), as well as domain-specific abilities in language in distributed frontal, temporal, and occipital areas (TFCE; *p*_FWE_ = .013; peak vertex = x: 49, y: -4, z: -9; Figure 3B), attention in right superior parietal cortex (TFCE; *p*_FWE_ = .043; peak vertex = x: 23, y: -54, z: 68; Figure 3B), and executive function in right fusiform/lingual cortex (TFCE; *p*_FWE_ = .047; peak vertex = x: 14, y: -54, z: -7; Figure 3B). Rhythmic neurophysiological slowing covaried only with attention abilities in right inferior frontal cortex (TFCE; *p*_FWE_ = .039; peak vertex = x: 50, y: 32, z: -13; Figure 3B). Mean neurophysiological slowing values per each region of the Yeo 7-networks atlas^74^ indicated greatest slowing in visual and dorsal attention networks, and weakest effects in somato-motor, ventral attention, and fronto-parietal networks (Figure S1). All the reported slowing effects and relationships to clinical metrics remained significant (all *p*’s < .005) after inclusion of confounds in the respective linear models, including head motion, eye movements, heart rate variability, and the number of epochs used per participant for analysis.

**Figure 3.**
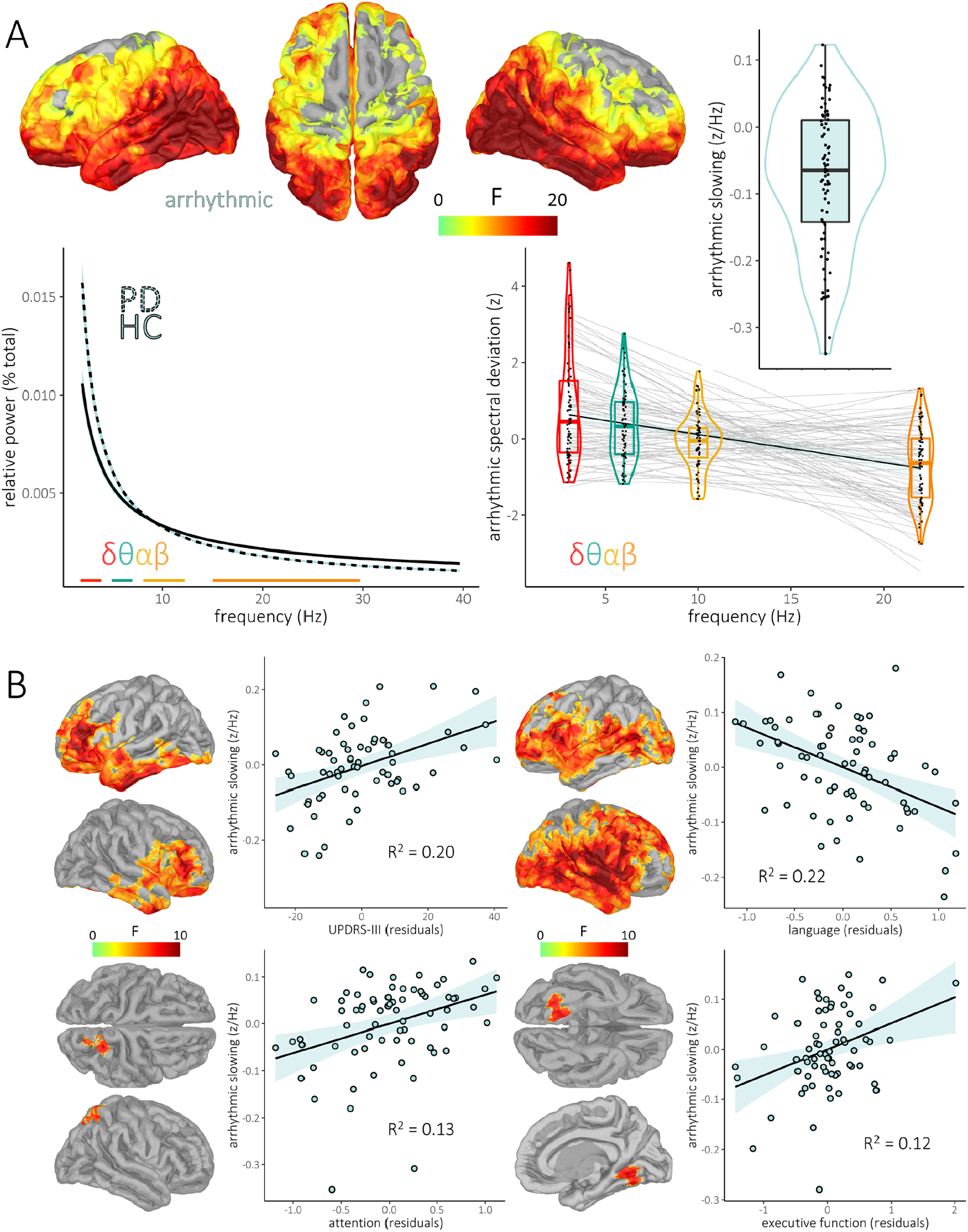
Arrhythmic neurophysiological slowing associated with clinical impairments in Parkinson’s disease. Similar to Figure 2, but with neurophysiological slowing computed using the arrhythmic (i.e., aperiodic) component of the parameterized spectra. (A) Cortical maps indicate significant clusters of arrhythmic neurophysiological slowing in patients with Parkinson’s disease (PD) after stringent multiple comparisons correction. Power spectra to the bottom left indicate the underlying data used to compute neurophysiological slowing from the cortical vertex exhibiting the strongest effect, with colored bars underneath showing the bandwidths of typical frequency-band definitions used for averaging. The plot to the bottom right shows the individual patient spectral deviations at this same cortical vertex for each frequency, with the light grey lines-of-best fit indicating individual neurophysiological slowing slopes, and the overlaid black line and blue shaded area representing the overall group effect and 95% confidence intervals, respectively. These individual and mean neurophysiological slowing effects are also represented as single dots in the scatterplot to the top right. (B) Cortical maps indicate significant clusters where neurophysiological slowing was associated with cognitive function in patients with PD. Associated scatterplots indicate the nature and strength of this relationship at the cortical vertex exhibiting the strongest effect, with lines-of-best-fit, 95% confidence intervals, and *R*^*2*^ values overlaid.

**Figure 4.**
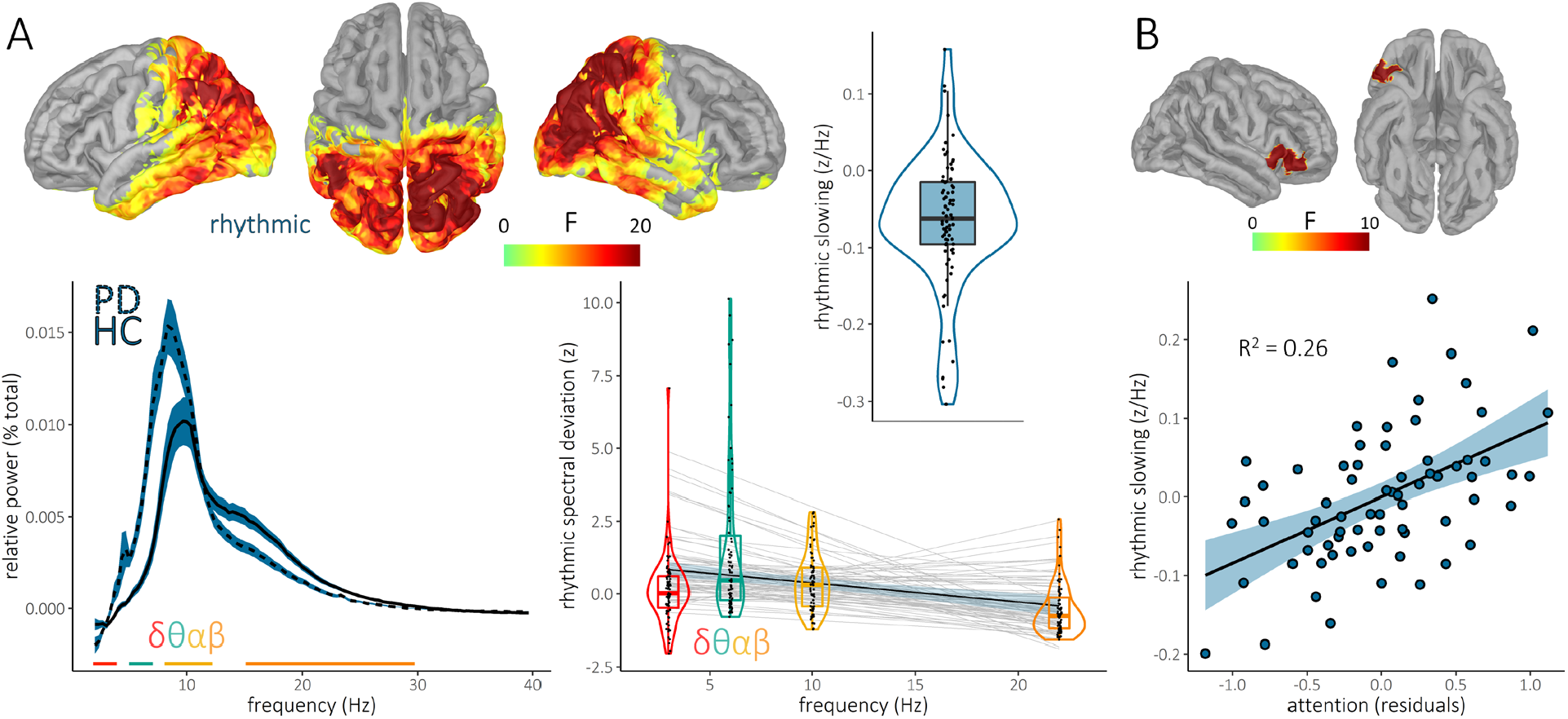
Rhythmic neurophysiological slowing associated with clinical impairments in Parkinson’s disease. Similar to Figure 2, but with neurophysiological slowing computed using the rhythmic (i.e., aperiodic-corrected) component of the parameterized spectra. (A) Cortical maps indicate significant clusters of rhythmic neurophysiological slowing in patients with Parkinson’s disease (PD) after stringent multiple comparisons correction. Power spectra to the bottom left indicate the underlying data used to compute neurophysiological slowing from the cortical vertex exhibiting the strongest effect, with colored bars underneath showing the bandwidths of typical frequency-band definitions used for averaging. The plot to the bottom right shows the individual patient spectral deviations at this same cortical vertex for each frequency band, with the light grey lines-of-best fit indicating individual neurophysiological slowing slopes, and the overlaid black line and blue shaded area representing the overall group effect and 95% confidence intervals, respectively. These individual and mean neurophysiological slowing effects are also represented as single dots in the scatterplot to the top right. (B) Cortical maps indicate the significant cluster where neurophysiological slowing was associated with attention function in patients with PD. Associated scatterplots indicate the nature and strength of this relationship at the cortical vertex exhibiting the strongest effect, with the line-of-best-fit, 95% confidence interval, and *R*^*2*^ value overlaid.

### Associations Between Neurophysiological Slowing and Clinical Impairments Exhibit a Spatial Gradient Across the Cortex

We observed that the nature of the relationships between cortical slowing and clinical impairments changed systematically across the sagittal plane of the cortex, with more posterior relationships generally indicating impairment (i.e., greater slowing associated with worse cognitive outcomes) and more anterior relationships indicating compensation. To test this effect empirically, we developed and implemented a new non-parametric method and found evidence of significant posterior – anterior (1,000 permutations; *b* = 3.57, *p* = .002) and superior – inferior (1,000 permutations; *b* = -5.64, *p* = .004) spatial gradients, such that stronger slowing in superior parietal cortices related to worse clinical impairments, while greater slowing in inferior frontal regions was associated with better preserved motor and cognitive functions (Figure 5A). These spatial gradient effects did not differ between the rhythmic and arrhythmic slowing models (1,000 permutations; posterior – anterior: *p* = .848; superior – inferior: *p* > .999; Figure 5B), and remained significant after correction for confounds (i.e., head motion, eye movements, and heart rate variability; 1,000 permutations; posterior – anterior: *p* < .001; superior – inferior: *p* = .014; Figure S2).

**Figure 5.**
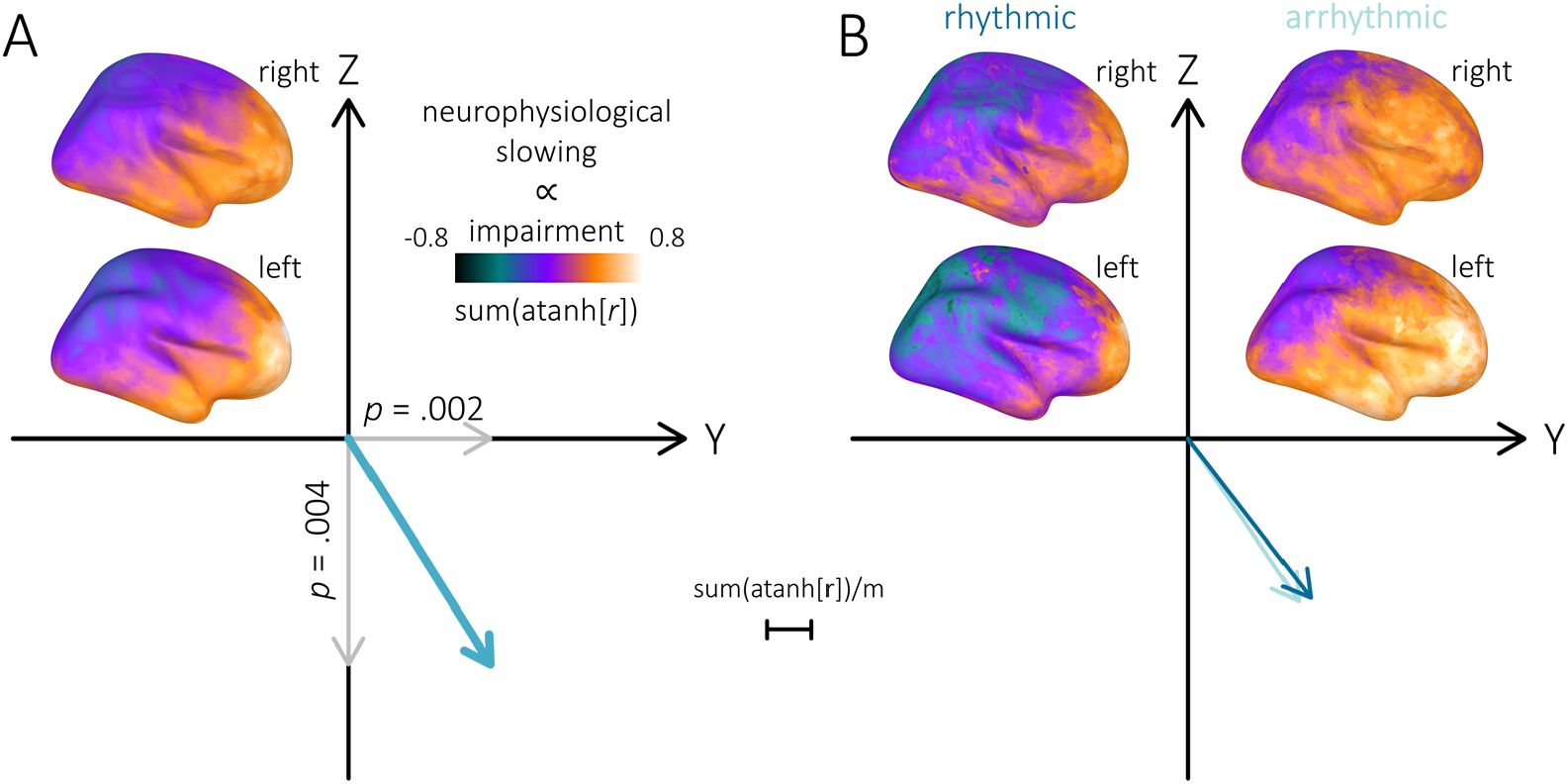
Anatomical gradient of clinical effects of neurophysiological slowing in Parkinson’s disease. (A) Cortical maps indicate the nature and strength of relationships between neurophysiological slowing and clinical impairments (i.e., partial correlations linearly-scaled and summed across motor and cognitive domains) along the cortex of patients with Parkinson’s disease, with lower values indicating a more pathological relationship (i.e., greater slowing predicts worse clinical deficits) and higher values indicating a possible compensatory effect. Grey vectors plotted along the cardinal anatomical axes are unstandardized beta weights from a multiple regression of the neurophysiological slowing – clinical impairment relationships on the relevant anatomical coordinates (X: left – right; Y: posterior – anterior; Z: inferior – superior), and indicate the magnitude and direction of the significant anatomical gradient effects. Overlaid p-values were generated using a non-parametric permutation approach and indicate statistical significance per each axis of the gradient effect. The blue vector indicates the magnitude and direction of the overall significant anatomical gradient effect. (B) Cortical maps again indicate the nature and strength of the neurophysiological slowing – clinical impairment relationships across the cortex of patients with Parkinson’s disease, but with neurophysiological slowing computed using the rhythmic (left) and arrhythmic (right) components of the parameterized spectra separately. The significant anatomical gradient effects observed in the non-parameterized neurophysiological slowing data (panel A) did not differ between the rhythmic and arrhythmic models.

This anatomical-neurophysiological gradient was significantly modulated by meaningful PD clinical factors (Figure 6). Patients who reported subjective cognitive complaints exhibited a weaker posterior – anterior gradient effect than those who did not (1,000 permutations; Δ*b* = -7.90, *p* = .010; Figure 6A). A similar effect of dopamine agonist use was also observed, with those patients taking dopamine agonists showing a weaker posterior – anterior gradient effect (1,000 permutations; Δ*b* = -5.56, *p* = .048; Figure 6B). Further, we discovered that the left – right gradient differed significantly based on the laterality of symptom onset (1,000 permutations; Δ*b* = -3.35, *p* = .006; Figure 6C), such that left-onset patients exhibited a bias toward compensatory effects of cortical slowing in the left hemisphere, while we observed the mirrored effect in right-onset patients. Post-hoc testing of significant clinical subgroup differences in these gradient effects indicated that the dopamine agonist (1,000 permutations; *p* = .042; Figure S2) and symptom laterality (1,000 permutations; *p* = .032; Figure S3) effects were specific to the model considering only motor impairments, while the effect of subjective cognitive complaints was specific to cognitive abilities (1,000 permutations; *p* = .006; Figure S4).

**Figure 6.**
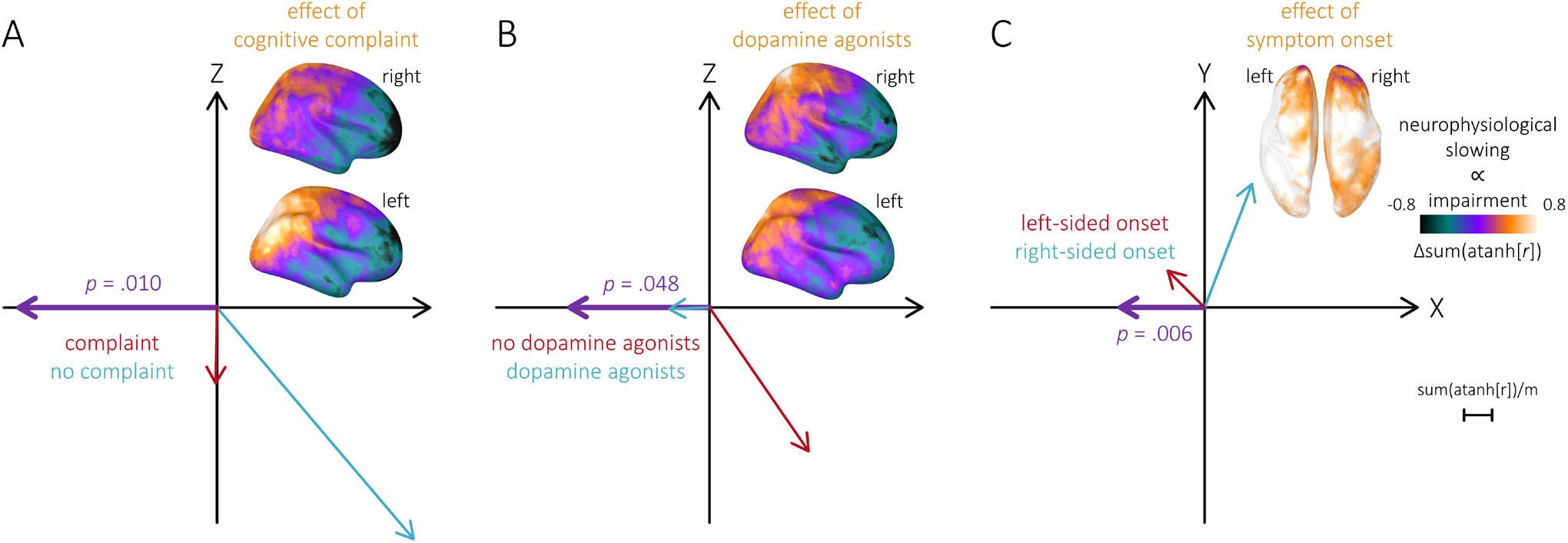
The anatomical gradients of clinical effects of neurophysiological slowing in Parkinson’s disease are clinically meaningful. Cortical maps indicate differences in the nature and strength of relationships between neurophysiological slowing and clinical impairments in patients with Parkinson’s disease, as a function of binary clinical features, including (A) the presence of subjective cognitive complaints, (B) the use of dopamine agonists, and (C) the laterality of initial symptom onset. Purple vectors plotted along the cardinal spatial axes are unstandardized beta weights from a multiple regression of the neurophysiological slowing – clinical impairment relationships on the relevant spatial coordinates (X: left – right; Y: posterior – anterior; Z: inferior – superior), subtracted between the two clinical feature subgroups. Overlaid p-values were generated using a non-parametric permutation approach and indicate statistical significance per each axis of the difference in the gradient effect. The blue and red vectors indicate the magnitude and direction of the overall anatomical gradient effects per each clinical feature subgroup.

### Clinical Effects of Cortical Slowing Selectively Co-Localize with Neurotransmitter Receptor Densities

To test for spatial associations between the observed anatomical-neurophysiological gradient and normative neurochemical systems, we adapted the non-parametric method described above to data from *neuromaps*^69^. The relationship between neurophysiological slowing and clinical impairments co-localized selectively with normative densities of dopamine, serotonin, GABA, and norepinephrine systems, but not with densities of acetylcholine and glutamate systems, nor with overall synaptic density (Figure 7). Specifically, all three measures of dopaminergic density related positively to the anatomical- neurophysiological gradient (1,000 permutations; D1: β = 0.38, *p*_FDR_ < .001; D2: β = 0.40, *p*_FDR_ = .024; DAT: β = 0.24, *p*_FDR_ = .024), such that regions with higher dopamine receptor/transporter density in health exhibited a compensatory effect of slowing in patients with PD. Similar positive associations were found for four of the six tested serotonergic density measures (1,000 permutations; 5-HT1a: β = 0.49, *p*_FDR_ = .023; 5- HT2a: β = 0.30, *p*_FDR_ = .017; 5-HT4: β = 0.43, *p*_FDR_ = .024; 5-HTT: β = 0.35, *p*_FDR_ = .024). In contrast, both GABAergic (1,000 permutations; GABAa: β = -0.34, *p*_FDR_ = .038) and noradrenergic (1,000 permutations; NET: β = -0.53, *p*_FDR_ = .038) densities related negatively to the gradient effect, such that regions with higher healthy receptor/transporter density exhibited a stronger pathological effect of cortical slowing in PD.

**Figure 7.**
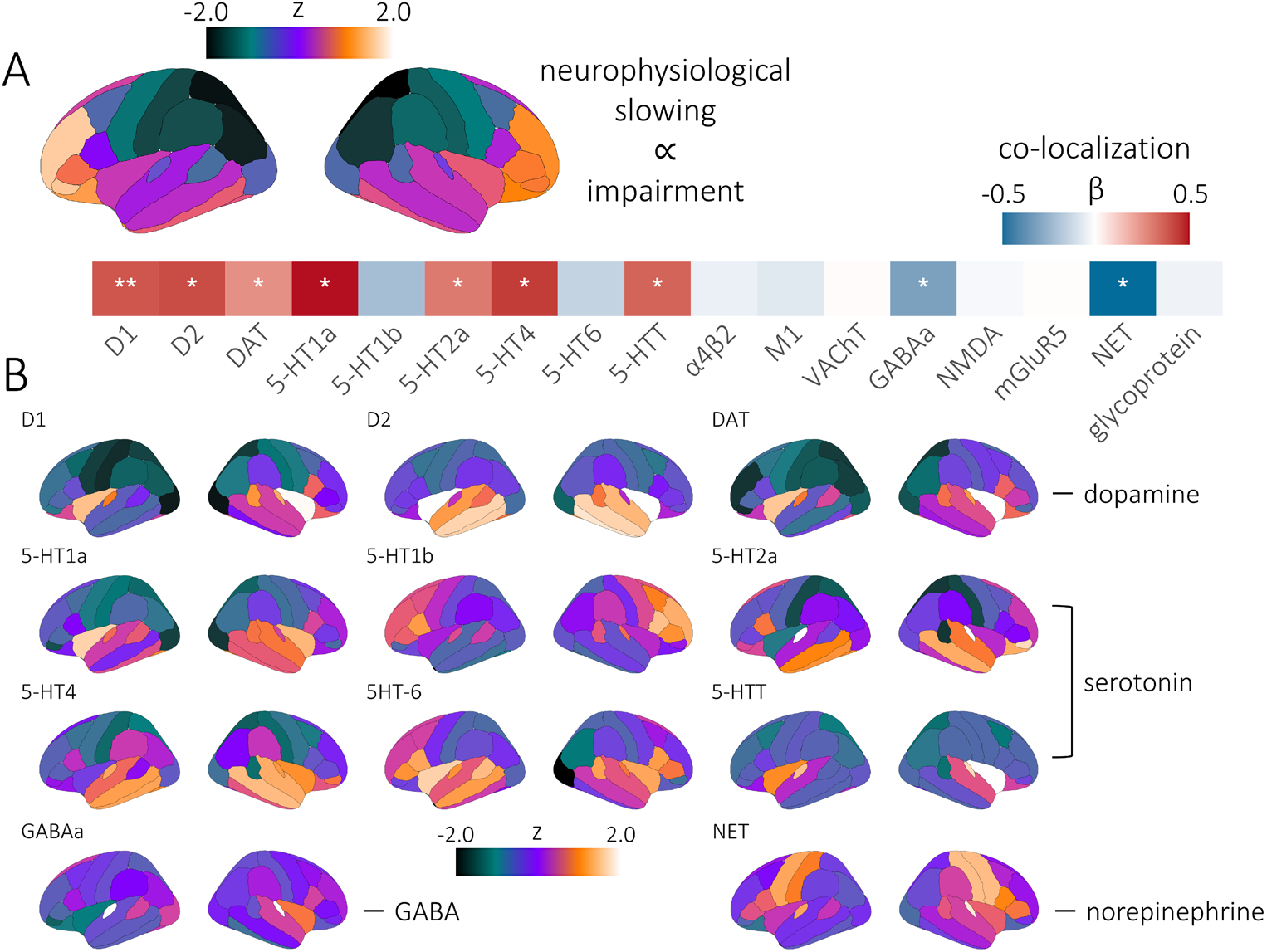
Clinical effects of neurophysiological slowing selectively co-localize with receptor densities. (A) Parcellated cortical maps indicate the nature and strength of relationships between neurophysiological slowing and clinical impairments (i.e., partial correlations linearly-scaled and summed across motor and cognitive domains, z-scored across brain regions) in patients with Parkinson’s disease. The vector heatmap below indicates the strength (standardized β) and statistical significance (\**p*_FDR_ < .05, \*\**p*_FDR_ < .005) of co-localization between the neurophysiological slowing-clinical relationship and each neuromap measure, including dopamine (D1, D2, and DAT), serotonin (5-HT1a, 5-HT1b, 5-HT2a, 5-HT4, 5- HT6, 5-HTT), acetylcholine (α4β2, M1, VAChT), GABA (GABAa), glutamate (NMDA, mGluR5), norepinephrine (NET), and synapse density (glycoprotein). (B) Parcellated cortical maps indicate the density of each neuromap measure, z-scored across brain regions.

### Inter-regional Functional Connectivity is Slowed in Parkinson’s Disease

Using the cortical location with the strongest neurophysiological slowing effect as a seed region (x: -50, y: -77, z: 1), we examined whether functional connectivity between this region and the rest of the cortex also exhibited a significant slowing effect in patients with PD. We found that inter-regional connections between the seed region and a widely distributed network of frontal, somato-motor, and superior parietal regions were significantly slowed in patients (TFCE; *p*_FWE_ < .001; peak vertex = x: 14, y: -61, z: 70; Figure S3). No significant relationships between this connectivity slowing effect and clinical outcomes were observed when stringent corrections were applied for multiple comparisons across cortical locations, but the magnitude of connectivity slowing at the peak of this effect did significantly relate to memory abilities (*t*(62) = 2.46, *p* = .017). Both the connectivity slowing main effect and its relationship to memory scores were robust to confounds (i.e., head motion, eye movements, heart rate variability, and the number of epochs used per participant for analysis; main effect: *p* < .001; memory relationship: *p* = .030).

## Discussion

After decades of literature suggesting a pathological shift in neurophysiological signal power from high to low frequencies in patients with neurodegenerative disorders^10-12,39-41,75-84^, recent advances in capturing these multi-spectral effects have documented their anatomical distribution and relevance to clinical features^19,42,52^. In the current work, we advance this line of research in patients with Parkinson’s disease using a marker of neurophysiological slowing that recently showed associations with cognitive impairments and amyloid proteinopathy in patients with Alzheimer’s disease^52^.

We find that patients with PD do exhibit broad neurophysiological slowing effects across posterior parietal, temporal, occipital, and inferior frontal cortices. This effect concerns both the rhythmic and arrhythmic components of the neurophysiological spectrum. Further, we show that the magnitude of this slowing effect is associated with individual cognitive and motor functions. Most notably, slowing across bilateral fronto-temporal cortical regions is related to better language abilities, while slowing in an ensemble of right-lateralized inferior frontal, somato-motor, and superior parietal regions is associated with worse attention scores. Rhythmic and arrhythmic slowing are differentially related to these observations: the relationship with language abilities was only recapitulated with arrhythmic slowing, while the relationship with attention was found in both the rhythmic and arrhythmic analyses, but with differing anatomical definitions. Arrhythmic slowing related to worse attention in right superior parietal regions, while rhythmic slowing exhibited the same association in right inferior frontal cortex. We also found relationships involving arrhythmic slowing that were not detected in the cortical slowing measures computed using the non- parameterized spectra, including a robust association between arrhythmic cortical slowing and better motor function (i.e., lower UPDRS-III scores) in bilateral prefrontal and anterior temporal cortices. Together, these results highlight not only the potential clinical relevance of cortical slowing in patients with PD, but also the insight gained by analyzing the respective effects of rhythmic and arrhythmic spectral features on neurophysiological slowing across patient populations.

At the macro-anatomical scale, we also show that the nature of these clinical-neurophysiological relationships varies across the cardinal sagittal axis of the brain. This anatomical-neurophysiological gradient indicates that slowing in superior and posterior cortices relates to worse clinical condition (i.e., higher UPDRS-III and lower neuropsychological scores), while slowing in inferior and anterior regions is associated with better motor and cognitive abilities, indicating compensation. We also found that this gradient is affected by key clinical factors: it is reduced by a dopamine agonist regimen, and it is stronger in patients with no subjective cognitive complaints. Further, although no overall left – right anatomical gradient was observed across all patients with PD, a marked difference in the gradient effect was observed along this axis when patients were sorted according to the laterality of their initial symptom onset, such that cortical slowing related to clinical compensation on the less affected hemisphere.

We identify four neurochemical systems as candidate contributors to this clinical-neurophysiological gradient effect. In essence, we find that brain regions with higher normative dopamine and serotonin and lower GABA and norepinephrine densities tend to exhibit a compensatory effect of cortical slowing in these patients. All four of these neurotransmitter systems are impacted by PD^85,86^. In particular, loss of cortical dopamine systems is a strong predictor of cognitive dysfunction in PD^87^. In combination with our finding that the sagittal gradient of cortical slowing is normalized by the use of dopaminergic agonists, we interpret the dopaminergic co-localization of this effect as further evidence that neurophysiological compensation in PD is largely necessitated by frontal dopamine dysfunction. It should be noted, however, that manipulation of primary dopamine medications (i.e., levodopa) was not possible in this study, warranting caution when interpreting the causal nature of these effects.

Taken together, these results suggest that the clinical impact of expressions of aberrant neurophysiological activity in PD is dependent on their anatomical locus and neurochemical basis: the same multi-spectral neurophysiological patterns indicating impairment in one brain region indicate compensation elsewhere on the cortex. These findings may explain the highly variable clinical outcomes of anatomically-targeted rhythmic modulation of frequency-specific neurophysiological activity^20-26^. In fact, many of these studies have targeted the primary motor cortices, which our results indicate as a point of anatomical inflection along the compensation-impairment axis. We argue that future studies aiming to ameliorate cognitive and motor symptoms in patients with PD should be anatomically selective, normalizing cortical slowing in posterior parietal cortices, and/or enhancing cortical slowing over inferior frontal regions. Spatial targeting of neuromodulation might also be personalized per patient. For example, distinct protocols may be advised depending on the laterality of symptom onset, prescription of dopaminergic agonists, and/or presence of subjective cognitive complaints.

To our knowledge, this is also the first report of both rhythmic and arrhythmic contributors to neurophysiological slowing effects in any patient population. Separating these contributions allowed us to detect relationships to motor function and cognition that were not significant when using non-parameterized neurophysiological spectra. This provides evidence that rhythmic and arrhythmic slowing effects are at least partially distinct. We also anticipate these findings will impact ongoing research on rhythmic neuromodulation for the treatment of patients with neurodegenerative disorders. Although we report clinically-relevant rhythmic components of cortical slowing, we also find that shifts in the arrhythmic spectra are associated with clinical features. We foresee that future research will investigate whether the cortical slowing effects reported previously in other patient groups^52,88,89^ exhibit similar distinctions between arrhythmic and rhythmic components of neurophysiological signals.

We also find that cortical slowing effects in PD are not confined to local changes in spectral power, but also affect frequency-specific inter-regional connectivity. The slowing effects of cortico-cortical functional connectivity reported herein may indicate a shift towards slower, more stable channels of neurophysiological communication in PD. However, this hypothesis needs to be tested directly in future work. We found that these connectivity slowing effects were widely distributed across the cortex, and argue that the data provide a proof-of-concept of the applicability of cortical slowing measures to other types of multi-spectral data (e.g., frequency-specific functional connectivity) and in other patient groups.

In sum, we show that patients with Parkinson’s disease exhibit neurophysiological slowing across multiple cortical regions, contributed by both rhythmic and arrhythmic spectral components. Cortical slowing is associated with worse motor function and cognition in superior parietal regions, but transitions to a compensatory effect along a superior – inferior and posterior – anterior anatomical gradient towards inferior frontal regions. This sagittal gradient effect indicates a need for more evidence-based targeting of neuromodulation therapies. We also demonstrate proof-of-concept for slowing of frequency-defined cortico-cortical functional connectivity in PD.

## Methods

### Participants

The Research Ethics Board at the Montreal Neurological Institute reviewed and approved this study. Written informed consent was obtained from every participant following detailed description of the study, and all research protocols complied with the Declaration of Helsinki. Exclusionary criteria for all participants included current neurological (other than PD) or psychiatric disorder; MEG contraindications; and unusable MEG or demographic data. All participants completed the same MEG protocols with the same instrument at the same site.

Patients with mild to moderate (Hoehn and Yahr scale: 1 – 3) idiopathic PD were enrolled in the Quebec Parkinson Network (QPN; https://rpq-qpn.ca/)^53^ initiative, which comprises extensive clinical, neuroimaging, neuropsychological, and biological profiling of each participant. A final sample of 79 participants with PD fulfilled the inclusion criteria. All patients with PD were prescribed a stable dosage of antiparkinsonian medication with satisfactory clinical response prior to study enrollment. Patients were instructed to take their medication as prescribed before research visits, and thus all data were collected in the practically-defined “ON” state.

Neuroimaging data from 65 healthy older adults were collated from the PREVENT-AD (N = 50)^54^ and Open MEG Archive (OMEGA; N = 15)^55^ data repositories to serve as a comparison group for the patients with PD. These participants were selected so that their demographic characteristics, including age (Mann-Whitney U test; *W* = 2349.50, *p* = .382), self-reported sex (chi-squared test; χ^2^ = 0.65, *p* = .422), handedness (chi-squared test; χ^2^ = 0.25, *p* = .883), and highest level of education (Mann-Whitney U test; *W* =2502.50, *p* = .444), did not significantly differ from those of the patient group. Group demographic summary statistics and comparisons, as well as clinical summary statistics for the patient group, are provided in Table 1.

**Table 1.** Group demographic comparisons and patient group clinical profile.

|  | Age<br>(years) | Sex<br>(% male) | Handedness<br>(# left/ambi) | Education<br>(years) |  |  |
| --- | --- | --- | --- | --- | --- | --- |
| HC (N = 65) | 63.02 (8.13) | 64.62 | 3/3 | 15.85 (3.72)* |  |  |
| PD (N = 79) | 64.60 (8.21) | 70.89 | 5/3 | 15.05 (3.04) |  |  |
| *p* | .382 | .422 | .883 | .444 |  |  |
|  | MoCA<br>(N = 70) | UPDRS-III<br>(N = 61) | Hoehn &<br>Yahr<br>(N = 57) | % Reporting<br>Cognitive<br>Complaints<br>(N = 55) | % Left<br>Symptom<br>Onset<br>(N = 66) | % Taking<br>DA Agonists<br>(N = 66) |
| PD |  |  |  |  |  |  |
| Range | 12 – 30 | 7 – 71 | 1 – 3 | - | - | - |
| Mean (SD) | 24.39 (3.98) | 32.44 (14.64) | 1.90 (0.70) | 58.18 | 51.52 | 31.82 |

HC: healthy control group; PD: Parkinson's disease group; MoCA: Montreal Cognitive Assessment; UPDRS-III: Unified Parkinson's Disease Rating Scale part III; DA: Dopamine. Unless otherwise indicated, values indicate means and associated parentheses indicate standard deviations. P-values indicate significance of between-groups Mann-Whitney U tests and chi-square tests for continuous and categorical variables, respectively. \*N = 59; highest level of education was not reported by six HC participants.

### Clinical & Neuropsychological Testing

Standard clinical assessments were available for most of the patients with PD, including data regarding gross motor impairment (Unified Parkinson’s Disease Rating Scale – part III [UPDRS-III]; N = 61)^56^, general cognitive function (Montreal Cognitive Assessment [MoCA]; N = 70)^57^, disease staging (Hoehn & Yahr scale; N = 57)^58,59^, symptom onset asymmetry (N = 66), use of dopamine agonists (N = 66), and subjective cognitive complaints (N = 55).

The patients were also asked to complete a series of detailed neuropsychological tests, with a final sample of 69 participants with PD providing useable data. These tests concerned five domains of cognitive function impacted in PD: attention (Digit Span – Forward, Backward, and Sequencing; Trail Making Test Part A), executive function (Trail Making Test Part B; Stroop Test – Colors, Words, and Interference; Brixton Spatial Anticipation Test), memory (Hopkins Verbal Learning Test-Revised [HVLT-R] – Learning Trials 1-3, Immediate and Delayed Recall; Rey Complex Figure Test [RCFT] – Immediate and Delayed Recall), language (Semantic Verbal Fluency – Animals & Actions; Phonemic Verbal Fluency – F, A & S; Boston Naming Test), and visuospatial function (Clock Drawing Test – Verbal Command & Copy Command; RCFT – Copy). To utilize as much available data as possible, missing values were excluded pairwise from analysis per each test. Negatively-scored test values were sign-inverted, the data for each individual test were standardized to the mean and standard deviation of the available sample, and these z-scores were then averaged within each domain listed above to derive domain-specific metrics of cognitive function. To corroborate the statistical independence of these domain composite scores, we computed a ratio of z-scores in the patient group representing the mean of all pairwise relationships (i.e., linearly-scaled Pearson correlation coefficients) amongst intra-domain tests, divided by the mean of all relationships with inter-domain tests. All domains had a ratio of z_intra_/z_inter_ > 1.50, and the mean z_intra_/z_inter_ ratio over all domains was 2.12 (SD = 0.30). This indicates that these domains were about twice more internally- than externally related on average. The mean across all five domains was also computed for each patient to represent general cognitive function. Importantly, as some participants were missing data on one or more tests within each domain, we verified that none of the domain scores were related to the number of tests used for their computation across individuals (attention: *r* = .04, *p* = .734; memory: *r* = -.19, *p* = .124; visuospatial function: no missing data; executive function: *r* = -.19, *p* = .117; language: *r* = -.08, *p* = .539; global function: *r* = -.14, *p* = .239; all BF_10_’s < 0.50). Demographically-corrected neuropsychological data were not available for this study, therefore, demographic factors significantly covarying with cognitive domain scores were included as nuisance covariates in all relevant statistical models. No significant impact of self-reported sex, highest level of education, nor handedness was found on any of the neuropsychological domain scores (all *p*’s > .20). In contrast, age was a moderate-to-strong predictor of neuropsychological testing performance (memory: *r* = -.34, *p* = .004; attention: *r* = -.22, *p* = .068; visuospatial function: *r* = -.54, *p* < .001; executive function: *r* = -.43, *p* < .001; language: *r* = -.26, *p* = .030). Accordingly, all statistical analyses utilizing these neuropsychological data included age as a nuisance covariate.

### Magnetoencephalography Data Collection and Analyses

Eyes-open resting-state MEG data were collected from each participant using a 275-channel whole-head CTF system (Port Coquitlam, British Columbia, Canada) at a sampling rate of 2400 Hz and with an antialiasing filter with a 600 Hz cut-off. Noise-cancellation was applied using CTF’s software-based built-in third-order spatial gradient noise filters. Recordings lasted a minimum of 5 min^60^ and were conducted with participants in the seated position as they fixated on a centrally-presented crosshair. The participants were monitored during data acquisition via real-time audio-video feeds from inside the MEG shielded room, and continuous head position was recorded during all sessions.

MEG preprocessing was performed with *Brainstorm*^61^ unless otherwise specified, with default parameters and following good-practice guidelines^62^. The data were bandpass filtered between 1–200 Hz to reduce slow-wave drift and high-frequency noise, and notch filters were applied at the line-in frequency and harmonics (i.e., 60, 120 & 180 Hz). Signal space projectors (SSPs) were derived around cardiac and eye- blink events detected from ECG and EOG channels using the automated procedure available in *Brainstorm*^63^, reviewed and manually-corrected where necessary, and applied to the data. Additional SSPs were also used to attenuate stereotyped artifacts on an individual basis. Artifact-reduced MEG data were then epoched into non-overlapping 6-second blocks and downsampled to 600 Hz. Data segments still containing major artifacts (e.g., SQUID jumps) were excluded for each session on the basis of the union of two standardized thresholds of ± 3 median absolute deviations from the median: one for signal amplitude and one for its numerical gradient. An average of 79.72 (SD = 13.82) epochs were used for further analysis (patients: 83.78 [SD = 7.24]; controls: 74.77 [SD = 17.82]). Empty-room recordings lasting at least 2 minutes were collected on or near the same day as the data recordings and were processed using the same pipeline, with the exception of the artifact SSPs, to model environmental noise statistics for source analysis.

MEG data were coregistered to each individual’s segmented T1-weighted MRI (*Freesurfer recon-all*)^64^ using approximately 100 digitized head points. For participants without useable MRI data (N = 14 patients with PD; N = 3 healthy older adults), a quasi-individualized anatomy was created and coregistered with *Brainstorm* to the MEG data, by warping the default *Freesurfer* anatomy to the participant’s head digitization points and anatomical landmarks^65^. Source imaging was performed per epoch using individually-fitted overlapping-spheres head models (15,000 cortical vertices, with current flows of unconstrained orientation) and dynamic statistical parametric mapping (dSPM). Noise covariance estimated from the previously-mentioned empty-room recordings were used for the computation of the dSPM maps.

### Analyses of Cortical & Functional Connectivity Slowing

Cortical slowing was assessed per patient with PD using a previously-validated method^52^, implemented as a linear model of spectral power deviations from healthy participants as a function of frequency (Figure 1A). This model is continuously-scaled, spatially-resolved, and unbiased by the natural differences in neurophysiological signal amplitude observed as a function of frequency. We computed vertex-wise estimates of power spectral density (PSD) from the source-imaged MEG data using Welch’s method (3-s time windows with 50% overlap) and normalized the resulting PSD estimates to the total power of the frequency spectrum. These PSD data were next averaged over all artifact-free 6-second epochs for each participant, and the PSD root-mean-squares (RMS) across the three unconstrained current flow orientations at each cortical vertex location was projected onto a template cortical surface (*FSAverage*) for comparison across participants.

To disentangle the slowing effects due to rhythmic versus arrhythmic cortical activity in patients with PD, we parameterized the PSDs with *specparam* (*Brainstorm* Matlab version; frequency range = 2–40 Hz; Gaussian peak model; peak width limits = 0.5 −12 Hz; maximum n peaks = 3; minimum peak height = 3 dB; proximity threshold = 2 standard deviations of the largest peak; fixed aperiodic; no guess weight)^43^ and extracted the exponent of arrhythmic spectral components. The arrhythmic components of the power spectra were the aperiodic outputs of *specparam* and the rhythmic (i.e., aperiodic-corrected) spectra were derived by subtracting these arrhythmic components from the original PSDs. The PSD components were then averaged over canonical frequency bands (delta: 2–4 Hz; theta: 5–7 Hz; alpha: 8–12 Hz; beta: 15–29 Hz)^63^. For each PD participant, the resulting PSD maps of spectrally-resolved estimates of neurophysiological signal power were normalized per frequency band to the mean and standard deviation of the comparable maps from the control group, resulting in cortical maps of PD spectral deviations from healthy variants. We then fit a linear model across the four frequency bands per each participant and cortical vertex location using the *polyfit* function in Matlab and extracted the model slope. This procedure yielded cortical maps of linear trends in spectral neurophysiological deviations per patient with PD. In these maps, cortical locations with flat slopes indicate locations of no substantial spectral change with respect to expected healthy variants, while more negative slopes indicate locations of stronger cortical slowing effects.

In addition to deriving maps of cortical slowing, we also used the source-imaged MEG data to investigate the potential for slowing of inter-regional functional connectivity in patients with PD. We extracted the first principal component from the three elementary source time series at each vertex location in each participant’s native space, and derived whole-cortex functional connectivity maps, using the cortical location with the strongest overall slowing effect (back-transformed into each participant’s native space) as the seed. We used orthogonalized amplitude envelope correlations (AEC)^66,67^ as the connectivity measure, based on the same frequency-band definitions used for the previously-described cortical slowing derivations. We estimated connectivity over each epoch and averaged the resulting AEC estimates across epochs, yielding a single AEC map per participant and frequency band. We projected these individual AEC maps onto the same template cortical surface (*FSAverage*) for group analyses, and used the previously-described procedure to derive spatially-resolved maps of functional connectivity slowing per patient with PD.

### Testing of Cortical Clinical-Gradient Effects

To ensure that gyrification of the pial surface did not bias our estimation of absolute distance between neighboring cortical locations, we applied a smoothing kernel to the template surface coordinate matrix using the *tess_smooth* function in Brainstorm (100% smoothing; i.e., smoothing factor of 1 with 46 iterations). We then used a two-step procedure to test for spatial gradients in the relationships between clinical impairments and neurophysiological slowing along the cortical surface (Figure 1B). We first modeled linear relationships at each cortical location between neurophysiological slowing and both motor (i.e., UPDRS-III) and cognitive (i.e., mean cognitive domain scores) impairments, beyond the effects of age, using the *partialcorr* function in Matlab. The resulting Pearson correlation coefficient values were then normalized using the Fisher transform (i.e., the inverse hyperbolic tangent; using the *atanh* function in Matlab), the neuropsychology correlations were sign-reversed for comparability with those computed from the UPDRS-III scores, and the two were summed at each location to generate cortical maps of the association between neurophysiological slowing and clinical impairments. In the second step, we fit a multiple regression model to these data using the *regress* function in Matlab, with the summed Fisher-transformed correlation coefficients as the dependent variable and the three cardinal axes of the template brain space (i.e., X: left – right, Y: posterior – anterior, and Z: inferior – superior) as the independent predictors. The unstandardized beta weights for each predictor were extracted from this model, representing the absolute change in the slowing–clinical impairment relationship (i.e., sum(atanh[*r*])) per unit distance (i.e., meters) across the cortex. Where relevant, we also performed post-hoc testing separately for the motor (i.e., UPDRS-III) and cognitive (i.e., mean cognitive domain scores) impairment data using the same procedure.

### Co-localization with Normative Atlases of Neurotransmitter Receptor Density

To determine the neurochemical systems that contribute to the observed cortical clinical-gradient effects, we adapted the two-step procedure described above, substituting as predictors region-wise estimates of normative neurotransmitter receptor/transporter density^68,69^ for the cardinal spatial axis data. Mean cortical receptor distribution maps of 16 different receptors and transporters from 6 neurotransmitter systems were computed as in previous work^68^ and parcellated using the Desikan-Killiany atlas^70^. These included dopamine (D1: 13 adults, [11C]SCH23390 PET; D2: 92, [11C]FLB-457, DAT: 174, [123I]-FP-CIT), serotonin (5-HT1a: 36, [11C]WAY-100635; 5-HT1b: 88, [11C]P943; 5-HT2a: 29, [11C]Cimbi-36; 5-HT4: 59, [11C]SB207145; 5-HT6: 30, [11C]GSK215083; 5-HTT: 100, [11C]DASB), acetylcholine (α4β2: 30, [18F]flubatine; M1: 24, [11C]LSN3172176; VAChT: 30, [18F]FEOBV), GABA (GABAa: 16, [11C]flumazenil), glutamate (NMDA: 29, [18F]GE-179; mGluR5: 123, [11C]ABP688), and norepinephrine (NET: 77, [11C]MRB). In addition, to test the importance of total synapse density, we also extracted a similar map of synaptic vesicle glycoprotein 2A (76, [11C]UCB-J)^71^. To facilitate comparison of the clinical-gradient effects to these normative maps, we parcellated the source-imaged MEG PSDs using the mean within each region of the same atlas^70^, and recomputed the neurophysiological slowing metric per atlas region. These slowing values were then related to cognitive and motor function (controlling for effects of age), normalized (and, for the cognitive relationships, sign-reversed), and summed using the same procedure described for the clinical- gradient analysis (step 1). To enable comparisons across neurotransmitter systems, the density and neurophysiological slowing data were each standardized (i.e., z-scored) across cortical regions. Linear regressions were then used to derive standardized beta-weights representing the co-localization of the cortical clinical-gradient effect with each normative neurotransmitter map (step 2).

### Testing for Potential Confounds

We investigated possible confound effects due to participant head motion, eye movements, and heart-rate variability. We extracted the head-position indicator, EOG, and ECG channel time series RMS, respectively. Alongside age and the number of trials used for analysis per participant, these derivations were included in *post hoc* statistical models to examine the robustness of the initial effect(s) of interest against potential confounds.

### Statistical Analyses

Participants with missing data were excluded pairwise per model. A threshold of *p* < .05 was used to indicate statistical significance, and all tests were performed two-tailed unless otherwise specified.

We derived statistical comparisons across the cortical maps produced, covarying out the effect of age, using *SPM12*. Initial tests used parametric general linear models, with secondary corrections of the resulting *F*- contrasts for multiple comparisons across cortical locations using Threshold-Free Cluster Enhancement (TFCE; E = 1.0, H = 2.0; 5,000 permutations)^72^. We applied a final cluster-wise threshold of *p*_FWE_ < .05 to determine statistical significance, and used the TFCE clusters at this threshold to mask the original statistical values (i.e., vertex-wise *F* values) for visualization. We extracted data from the cortical location exhibiting the strongest statistical relationship in each cluster (i.e., the “peak vertex”) for subsequent analysis and visualization. Where appropriate, linear models were fit to these extracted data using the *lm* function in *R*^73^.

A non-parametric permutation approach was used to determine the statistical significance of the spatial gradient and neurotransmitter co-localization effects, wherein at each vertex the patient cortical slowing data were randomly permuted (using the *randperm* function in Matlab) and used to compute the partial correlations (step 1) and regressions (step 2). This process was repeated 1,000 times, and the resulting beta-weights were extracted to generate null distributions for each predictor. The original beta coefficients were then compared with their respective null distributions to generate non-parametric p-values. To test for significant subgroup effects on the spatial gradients based on binary clinical factors (i.e., subjective cognitive complaints, symptom onset laterality, and dopamine agonist use), we implemented the same two-step approach, using instead the difference in beta-weights (from step 2) between clinical groups as the statistic of interest.

### Data & Code Availability

Data used in the preparation of this work are available through the Clinical Biospecimen Imaging and Genetic (C-BIG) repository (https://www.mcgill.ca/neuro/open-science/c-big-repository)^53^, the PREVENT- AD open resource (https://openpreventad.loris.ca/)^54^, and the OMEGA repository (https://www.mcgill.ca/bic/resources/omega)^55^. Normative neurotransmitter density data are available from *neuromaps* (https://github.com/netneurolab/neuromaps)^69^. Code for MEG preprocessing and the neurophysiological slowing and spatial gradient analyses is available at https://github.com/aiwiesman/QPN_Slowing. Rejection of epochs containing artifacts was performed with the *ArtifactScanTool* (https://github.com/nichrishayes/ArtifactScanTool).

## Data Availability

Data used in the preparation of this work are available through the Clinical Biospecimen Imaging and Genetic (C-BIG) repository (https://www.mcgill.ca/neuro/open-science/c-big-repository), the PREVENT-AD open resource (https://openpreventad.loris.ca/), and the OMEGA repository (https://www.mcgill.ca/bic/resources/omega). Normative neurotransmitter density data are available from neuromaps (https://github.com/netneurolab/neuromaps). Code for MEG preprocessing and the neurophysiological slowing and spatial gradient analyses is available at https://github.com/aiwiesman/QPN_Slowing. Rejection of epochs containing artifacts was performed with the ArtifactScanTool (https://github.com/nichrishayes/ArtifactScanTool).

https://openpreventad.loris.ca/

https://www.mcgill.ca/bic/resources/omega

https://github.com/netneurolab/neuromaps

https://github.com/aiwiesman/QPN_Slowing

## Acknowledgments

This work was supported by grant F32-NS119375 to AIW from the United States National Institutes of Health (NIH); to JDSC as a doctoral fellowship from Natural Science and Engineering Research Council of Canada (NSERC); to EAF as a Foundation Grant from the Canadian Institutes of Health Research (CIHR; FDN- 154301) and the CIHR Canada Research Chair (Tier 1) of Parkinson’s Disease; and to SB from by a NSERC Discovery grant, the Healthy Brains for Healthy Lives initiative of McGill University under the Canada First Research Excellence Fund, the CIHR Canada Research Chair (Tier 1) of Neural Dynamics of Brain Systems and the NIH (1R01EB026299). Data collection and sharing for this project was provided by the Quebec Parkinson Network (QPN), the Pre-symptomatic Evaluation of Novel or Experimental Treatments for Alzheimer’s Disease (PREVENT-AD; release 6.0) program, and the Open MEG Archives (OMEGA). The funders had no role in study design, data collection and analysis, decision to publish, or preparation of the manuscript.

The QPN is funded by a grant from Fonds de recherche du Québec - Santé (FRQS). PREVENT-AD was launched in 2011 as a $13.5 million, 7-year public-private partnership using funds provided by McGill University, the FRQS, an unrestricted research grant from Pfizer Canada, the Levesque Foundation, the Douglas Hospital Research Centre and Foundation, the Government of Canada, and the Canada Fund for Innovation. Private sector contributions are facilitated by the Development Office of the McGill University Faculty of Medicine and by the Douglas Hospital Research Centre Foundation (http://www.douglas.qc.ca/). OMEGA and the Brainstorm app are supported by funding to SB from the NIH (R01-EB026299), a Discovery grant from the Natural Science and Engineering Research Council of Canada (436355-13), the CIHR Canada research Chair in Neural Dynamics of Brain Systems, the Brain Canada Foundation with support from Health Canada, and the Innovative Ideas program from the Canada First Research Excellence Fund, awarded to McGill University for the HBHL initiative.

## Notes

### Competing Interest Statement

The authors have declared no competing interest.

### Author Declarations

The Research Ethics Board at the Montreal Neurological Institute reviewed and approved this study. Written informed consent was obtained from every participant following detailed description of the study, and all research protocols complied with the Declaration of Helsinki.

