## Supplemental Figures for "A sagittal gradient of pathological and compensatory effects of neurophysiological slowing in Parkinson’s disease"

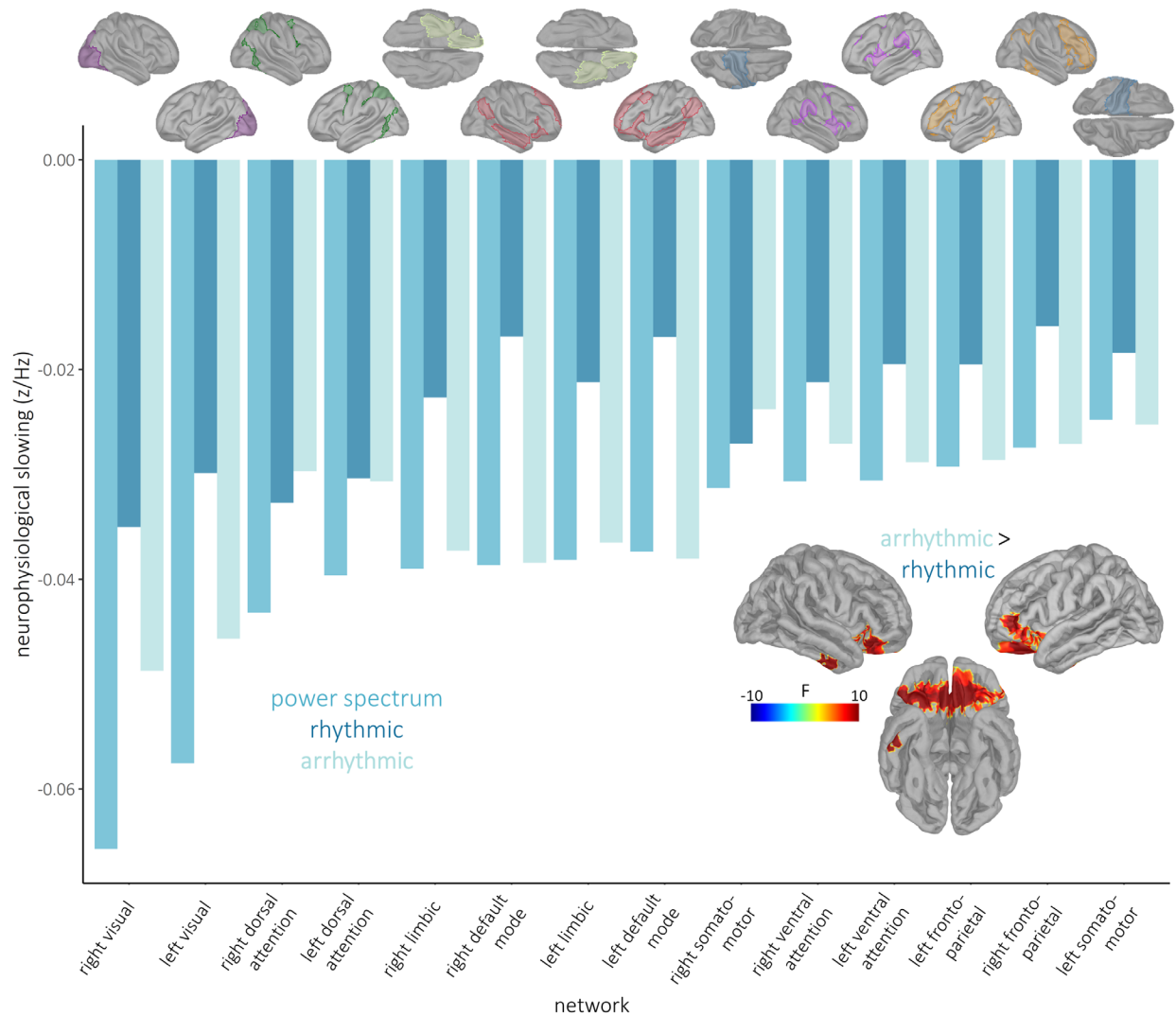

**Figure S1. Differences in arrhythmic versus rhythmic neurophysiological slowing and magnitude of the neurophysiological slowing effects across functional networks.** Bars indicate the mean neurophysiological slowing effect (y-axis) per each model (i.e., non-parameterized, rhythmic, and arrhythmic), averaged within each network of the Yeo 7-networks atlas<sup>74</sup>. Cortical maps above indicate the spatial extents of the networks used for averaging, with corresponding network labels on the x-axis. Inlaid cortical maps to the bottom right indicate the significant cluster where arrhythmic neurophysiological slowing was found to be stronger than rhythmic neurophysiological slowing.

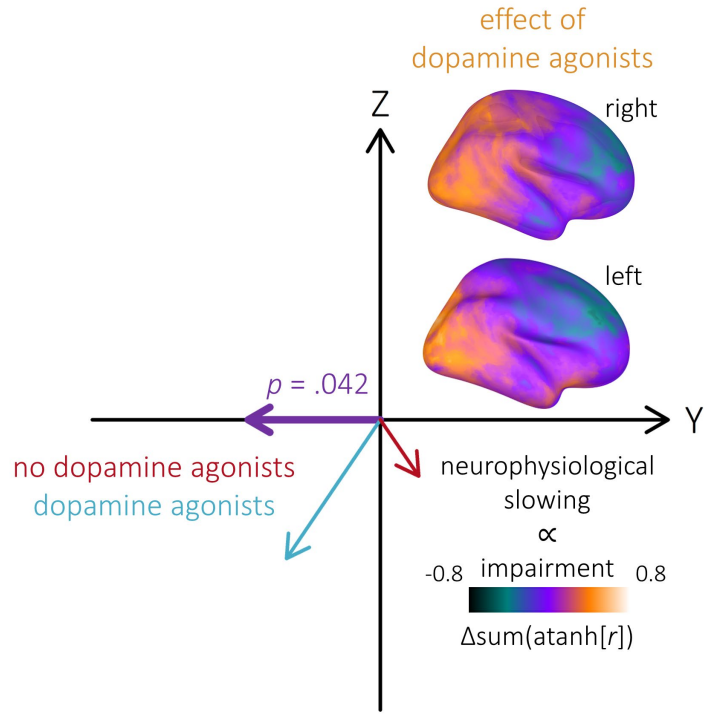

**Figure S2. Effect of dopamine agonist use on clinical – neurophysiological slowing gradients is specific to motor impairments.** Similar to Figure 6B, but considering only motor impairments (i.e., UPDRS-III data) in the computation of the anatomical gradient effects. No significant effect of dopamine agonist use on the clinical – neurophysiological slowing spatial gradient was observed when only cognitive impairments (i.e., averaged cognitive domain scores) were considered.

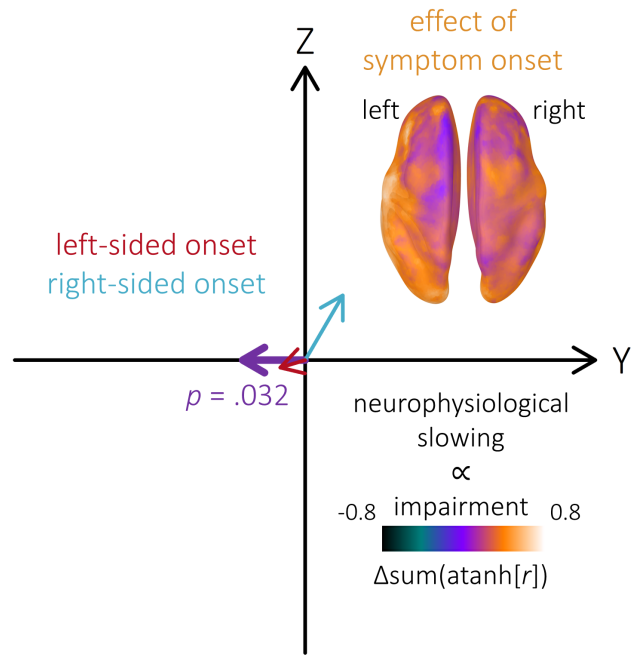

**Figure S3. Effect of symptom onset laterality on clinical – neurophysiological slowing gradients is specific to motor impairments.** Similar to Figure 6C, but considering only motor impairments (i.e., UPDRS-III data) in the computation of the anatomical gradient effects. No significant effect of symptom onset laterality on the clinical – neurophysiological slowing spatial gradient was observed when only cognitive impairments (i.e., averaged cognitive domain scores) were considered.

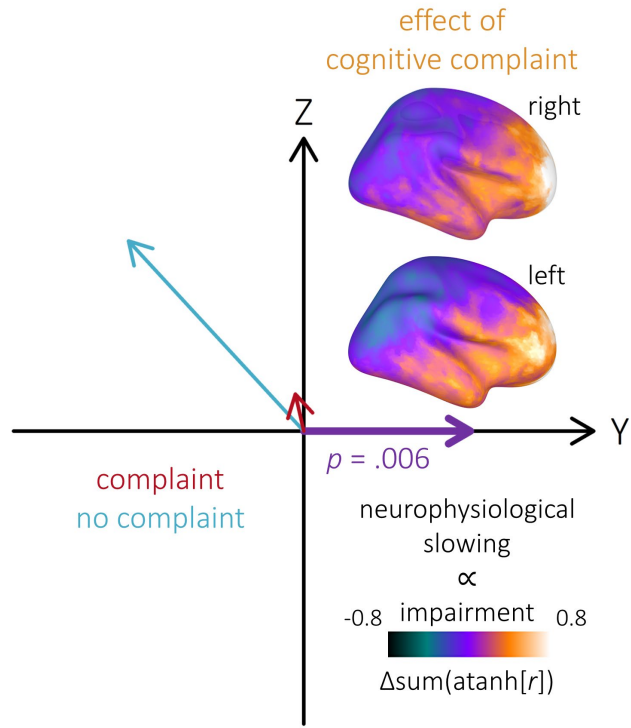

**Figure S4. Effect of subjective cognitive complaints on clinical – neurophysiological slowing gradients is specific to cognitive impairments.** Similar to Figure 6A, but considering only cognitive impairments (i.e., averaged cognitive domain scores) in the computation of the anatomical gradient effects. No significant effect of symptom onset laterality on the clinical – neurophysiological slowing spatial gradient was observed when only motor impairments (i.e., UPDRS-III data) were considered.

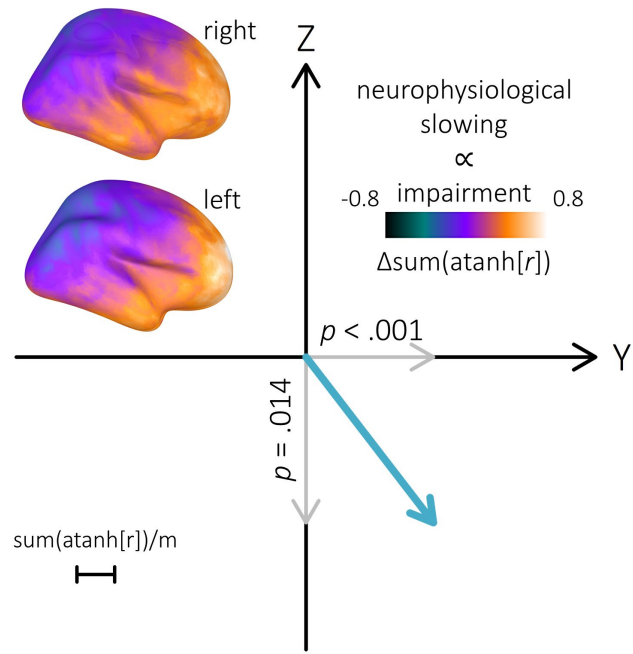

**Figure S5. Clinical – neurophysiological slowing spatial gradients are unaffected by confounds.** Comparable to Figure 5A, but with neurophysiological slowing – clinical impairment relationships modeled with inter-participant variability in head motion, eye movements, and heart rate as nuisance covariates. Both the posterior-anterior and superior-inferior anatomical gradients remained significant when controlling for these potential confounds.

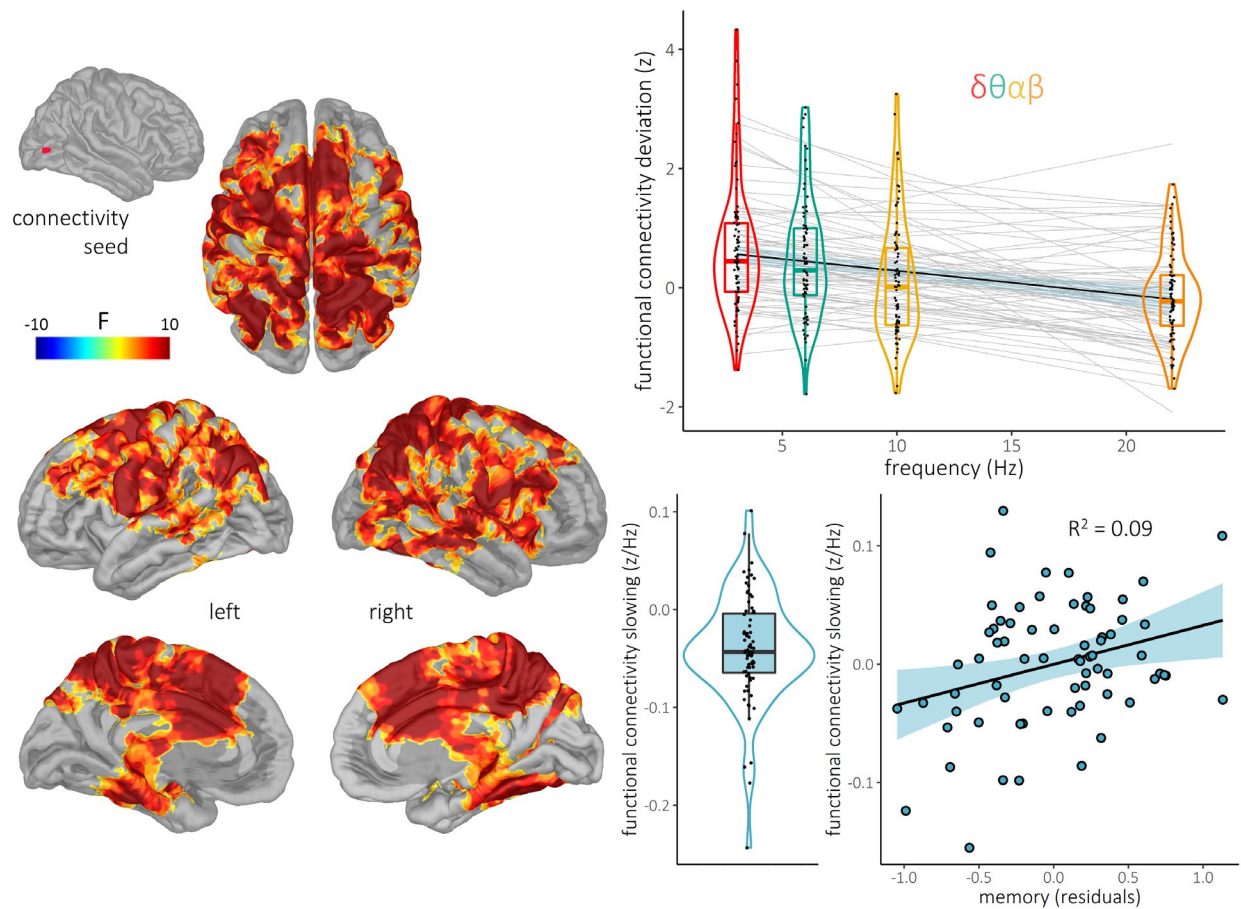

**Figure S6. Inter-regional functional connectivity is slowed in Parkinson's disease.** The cortical map to the top left indicates the location of the seed vertex (taken from the peak of the neurophysiological slowing main effect shown in Figure 2A) used for estimation of functional connectivity with the rest of the cortex. Statistical cortical maps to the left indicate the strength of slowing of functional connectivity between this seed and the rest of the cortex in patients with Parkinson's disease, relative to healthy adults. The plot to the top right shows the individual patient spectral deviations per frequency band from the vertex exhibiting the strongest connectivity slowing effect, with the light grey lines-of-best fit indicating individual connectivity slowing slopes, and the overlaid black line and blue shaded area representing the overall group effect and 95% confidence intervals, respectively. These individual and mean neurophysiological slowing effects are also represented as single dots in the scatterplot to the bottom middle. The scatterplot to the bottom right indicates the nature and strength of the relationship between connectivity slowing at the cortical vertex showing the strongest such effect and memory performance in patients with Parkinson's disease, with the line-of-best-fit, 95% confidence interval, and $R^2$ value overlaid.
